# Enhancing national cholera surveillance using rapid diagnostic tests (RDTs): a mixed methods evaluation

**DOI:** 10.1101/2025.01.10.25320331

**Authors:** ET Baumgartner, KN Williams, E Rai, EN Rosser, RP Marasini, S Dahal, A Shakya, J Lynch, K Karki, D Bajracharya, DA Sack, AK Debes

## Abstract

**Background:** Cholera rapid diagnostic tests (RDTs) can strengthen existing surveillance systems by offering a cost-effective screening method that improves understanding of cholera burden allowing for targeted prevention and control efforts. The RDT Implementation Strategy and Evaluation (RISE) project is the pilot study for Gavi’s innovative Diagnostic Procurement Platform which provides cholera RDTs to enhance national surveillance.

**Methods:** Implementation of cholera RDTs was evaluated following their distribution in 2023 to facilities within Nepal’s Early Warning and Reporting System (EWARS). Quantitative data was collected through EWARS surveillance reports, national-level and individual-level REDCap surveys from select facilities in Kathmandu. Key-informant interviews were also conducted in Kathmandu with personnel involved in cholera surveillance and response. Interviews were conducted using a semi-structured interview guide and analyzed according to inductively identified themes.

**Results:** Qualitative findings indicated generally positive perceptions of cholera RDTs, highlighting their speed and ease of use, and suitability for deployment in under-resourced areas by unskilled personnel. However, a lack of awareness of the RDTs, limited training, and concerns about the RDTs’ quality, availability, and costs were challenges raised consistently. Quantitative findings revealed underreporting of acute gastroenteritis (AGE) and cholera in EWARS and an underutilization of the cholera RDTs, with only 2.6% of reported AGE cases screened using an RDT.

**Discussion:** This field evaluation demonstrated that RDTs can have an important role in cholera surveillance but highlighted significant challenges with cholera lab capacity, reporting, and training. Both the qualitative and quantitative findings showed gaps in surveillance reporting, which were exacerbated by the complexity of adding RDTs without strong guidance as well as beliefs about the RDTs’ poor validity. These misconceptions and challenges need to be addressed at the local and national level to successfully scale-up cholera RDTs in Nepal and beyond.

## INTRODUCTION

Cholera is a highly contagious, diarrheal illness caused by the *Vibrio cholerae* O1 bacteria. [1]. Globally, an estimated 1.3 to 4.0 million cases and 21,000 to 143,000 deaths are attributed to cholera annually [2, 3, 4]. However, the World Health Organization (WHO) acknowledges that only 5%–10% of cholera cases are reported worldwide [2, 3] therefore, recent efforts to improve understanding of disease burden have included the incorporation of rapid diagnostic testing on a broader scale.

Logistics and cost constraints associated with microbiology testing are a major hinderance to accurate surveillance reporting. The gold standards for cholera diagnosis—microbial culture and PCR—have challenges including lengthy testing times, skilled laboratory staff requirements, and high costs [1, 8, 9, 10]. These drawbacks result in delays in outbreak confirmation and control, which leads to higher morbidity and mortality. Inaccuracies and delays in cholera reporting prevent precise disease burden estimates and create health and economic strain in affected countries [3, 4, 10, 11, 12, 13]. Without a comprehensive understanding of the disease’s geographic and temporal patterns, public health officials struggle to target interventions effectively, especially given the limited supply of the oral cholera vaccine (OCV) compared to the global at-risk population [14].

Cholera rapid diagnostic tests (RDTs) detect *V. cholerae* O1 and/or O139-specific antigens within 15 to 30 minutes [3, 9, 11, 15]. The UNICEF catalogue currently lists two cholera RDTs that are under review for pre-qualification, which includes an assessment of the quality, safety and efficacy by the WHO [14]. These RDTs can be administered by minimally trained staff at the bedside of a patient, requiring no cold chain or laboratory equipment [5, 15]. They decrease the need for confirmatory tests by serving as screening tools with a very high negative predictive value, especially in areas with low prevalence [9]. The RDTs are not recommended for use in determining treatment of patients, rather are meant for epidemiological surveillance only. Their timeliness and cost-effectiveness have the potential to facilitate improved surveillance, early outbreak detection, and more efficient responses to control the spread of bacteria [5]. Early field work has shown that the increasing utilization of cholera RDTs has facilitated cholera outbreak detection, particularly in endemic areas [6, 4, 18].

Gavi, The Vaccine Alliance established the Gavi-approved Diagnostics Initiative to support the procurement of cholera RDTs. The intention is to strengthen cholera surveillance through widespread distribution of cholera RDTs, and ultimately aid in targeting OCV campaigns to persons at greatest risk. Although some qualitative investigations have been conducted to understand uptake of OCV by patients and healthcare policy makers and providers, there remains a gap in qualitative investigations to understand stakeholder perceptions of cholera RDTs [25, 26]. Given that political support of cholera RDTs and provider willingness to use them are essential to achieve their potential benefits, understanding positive and negative perceptions of key stakeholders related to cholera RDTs is essential to ensure that such concerns and recommendations could be addressed and incorporated into national programs to increase cholera RDT uptake and impact. Our RDT Implementation Strategy and Evaluation (RISE) project seeks to fill this gap by understanding initial perceptions and usage of cholera RDTs following national RDT distribution in Nepal, with the goal of uncovering strategies that could be incorporated into a government-led national cholera RDT distribution program.

## METHODS

### Study site

Cholera outbreaks are reported in Nepal annually, often during the monsoon season which lasts from the end of May until September. During this time of year, frequent flooding and landslides interfere with water and sanitation infrastructure, making the spread of this pathogen more likely [7]. Electronic cholera surveillance is conducted using the Early Warning and Reporting System (EWARS), consisting of 118 sentinel hospitals that report on high priority diseases including cholera and acute gastroenteritis (AGE). According to the country’s most recent National Cholera Control Plan (NCCP), when there is a case of suspected cholera or a cluster of more than five AGE cases, EWARS sites must report within 24 hours so an outbreak investigation can be initiated in collaboration with the National Public Health Laboratory (NPHL). Cary Blair (CB) media is used to preserve samples from sites with potential outbreaks, which can be culture or PCR confirmed in the NPHL.

### Study design

The RDT Implementation Strategy and Evaluation (RISE) study in Nepal distributed RDTs to 114 of the 118 EWARS health facilities across Nepal in 2023. Facilities received between 10-50 RDTs, based on the average number of AGE cases seen at each facility; RDT shipments to 4 facilities could not be completed given logistical challenges. The goal of our study is to evaluate the implementation of RDTs in the EWARS system comparing national level cholera surveillance to an intensive cholera surveillance project led by International Vaccine Institute (IVI) in Kathmandu Valley (data not yet published). The RISE study design is depicted visually in Figure 1. Facilities participating in the intensive surveillance study received compensation for enrolling study participants, using the RDTs and completing the research forms. The study teams received comprehensive cholera RDT trainings.

**Figure 1.**
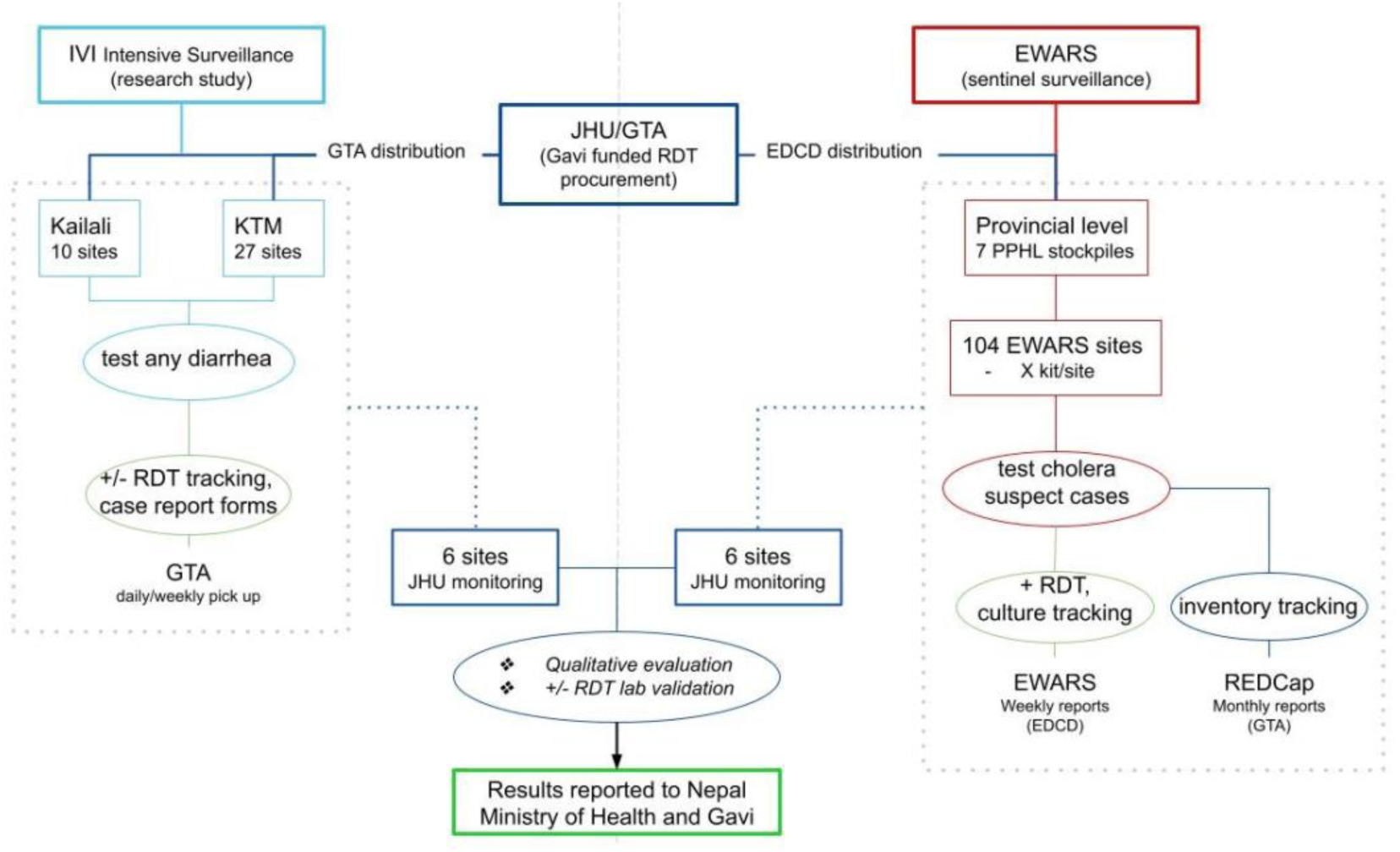
Flow chart depicting the RISE project operational and data collection activities.

The RISE project utilized a mixed methods approach to evaluate the RDT implementation in 2023: 1) a monthly cholera RDT utilization survey emailed to all 114 EWARS facilities that received RDTs through RISE, 2) individual level data collection at 12 EWARS facilities in Kathmandu Valley, and 3) qualitative interviews. The monthly survey included questions on number of cholera RDTs in stock, number of RDTs used, and number of positive RDTs. Sites participating in individual level data collection (which included 6 health facilities participating in the IVI’s intensive surveillance study and 6 facilities that were supported through the EWARS national surveillance system directed by the Epidemiology and Disease Control Division (EDCD)) completed an individual RDT survey each time an RDT was used. The survey included questions on patient age and sex, RDT result, confirmatory testing done, patient symptoms, antibiotic use, final diagnosis, and reporting to EWARS. All used RDTs were preserved for PCR testing to confirm the result [18]. We evaluated the use and implementation of cholera RDTs in the national EWARS sentinel sites compared to sites receiving intensive research-based surveillance through IVI.

Selection of the 12 nested surveillance sites was based on the number of AGE cases treated at the facility, its location relative to Kathmandu district, and their enrollment status in RISE or the intensive surveillance study. Qualitative interviews were conducted with clinicians and laboratory technicians from the 12 nested surveillance sites, as well as with key governmental stakeholders involved in cholera RDT implementation.

### Quantitative methods

Quantitative data for RISE was collected using REDCap electronic data capture tools hosted at Johns Hopkins University [18]. We also obtained national-level data from EWARS monthly surveillance reports for June through December 2023 from EDCD. All data was analyzed using R Studio version 2024.9.0.375 [19].

Monthly EWARS surveillance reports were used to calculate the total number of AGE and cholera events and the percent of cases for which a confirmatory lab test and/or RDT were performed. Results from the monthly cholera RDT utilization surveys and individual level REDCap surveys administered in the 12 RISE nested sites were used to calculate the frequency of RDT usage at EWARS facilities on the national and nested level, respectively.

### Qualitative Methods

From August through September 2023, qualitative key informant interviews were conducted with public health stakeholders and health personnel staff at the local health facilities. We used a purposive and snowball sampling strategy to identify laboratory technicians and physicians at the 12 nested data collection facilities in Kathmandu Valley, and government officials from the EDCD and NPHL Inclusion criteria comprised public health professionals and health workers who used cholera RDTs provided by the RISE Project, worked at sites where cholera RDTs were provided, or were involved in their distribution, implementation or analysis. These included physicians ordering the RDTs for their suspected cholera patients, laboratory technicians using the RDTs and verifying their results with further diagnostic testing, and government officials involved in RDT policy and implementation. Exclusion criteria precluded any person that had not used or coordinated the use of RDTs in a professional capacity, was under 18 years of age, or was not willing to give consent.

Interviews lasted 30 to 60 minutes, using semi-structured interview guides specific to each target group. Each interview was given a unique label, and all identifiers were removed prior to transcription. The interviewers took field notes and recorded their initial thoughts and impressions during and after each interview, reflecting on their positionality and noting any potential biases. When permission was granted, the interviews were recorded using a handheld device. Recordings were transcribed in Nepali and then translated into English.

Data analysis was an iterative process that involved a mixture of deductive and inductive coding [18]. The research team developed a set of deductive codes based upon the interview guide shown in Table 1 and incorporated additional emerging themes as the analysis progressed. Final codes are reported in Tables 1, 2, and 3 of the supplemental materials. Interview transcripts and expanded interview notes were uploaded into a coding framework in Excel, which was used to identify relevant quotes within each theme. To organize the findings, a thematic framework matrix in Excel was used to compare phenomena, identify emerging themes, and quotes into corresponding domains to inform a broader narrative.

**Table 1.**
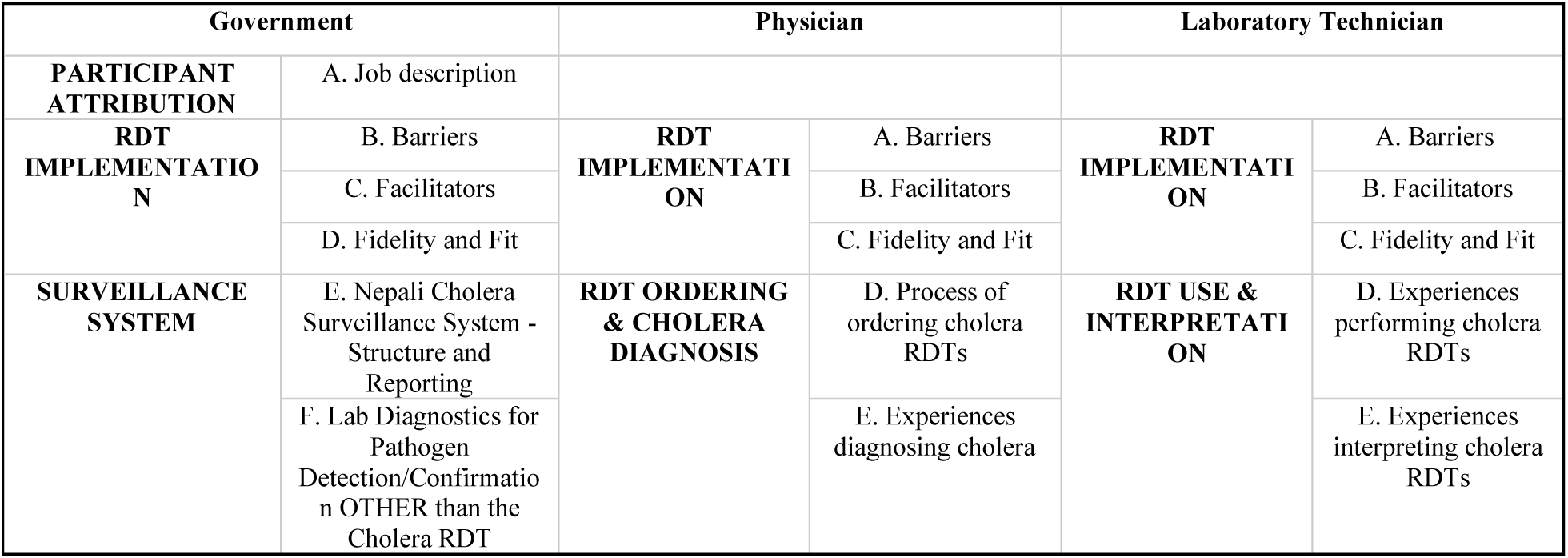
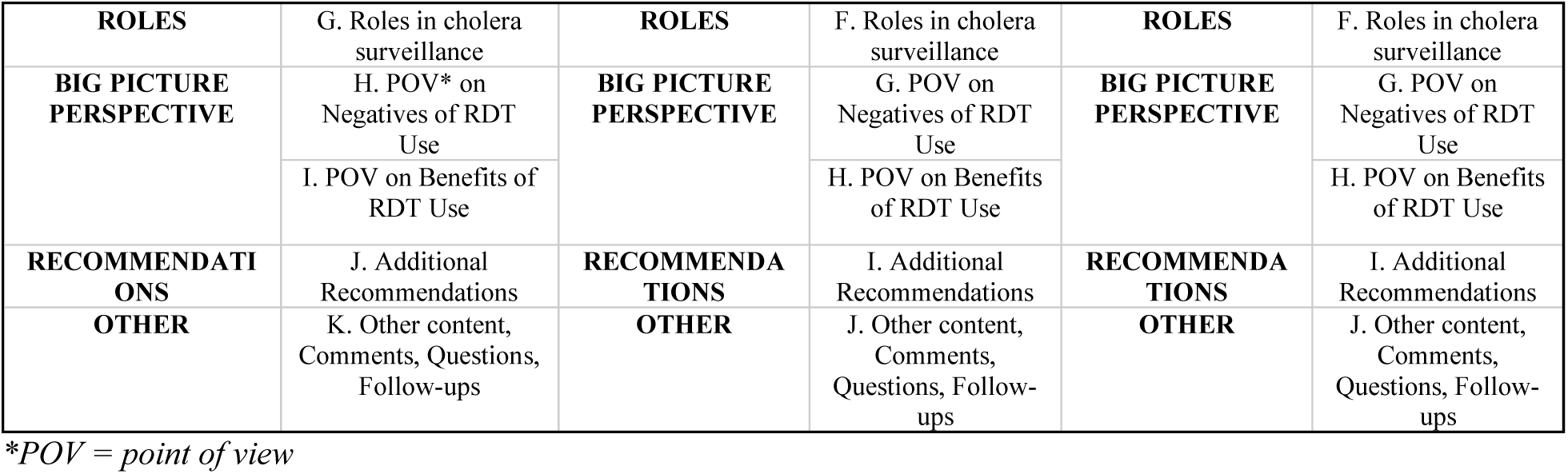
Guiding themes used during open coding analysis of interviews.

**Table 2.** Exemplary participant quotes categorized by topic.

| Theme | Speaker | Quote |  |
| --- | --- | --- | --- |
| RDTs facilitate cholera surveillance | Physician, teaching hospital | “It seems that the RDT seems to be sensitive one, but the specificity is a little bit low. So, for screening purpose, RDT test is easier and quick. So, initial test with RDT, if that comes positive, then confirmatory test is required, I think. And that can be sent to the NPHL or any center, nearby center. That will be useful.” | A1 |
| RDTs are easy to perform | Lab technician, private hospital | “Because in the summer season, the diarrhea cases are increasing. So, to do the microscopy for the cholera, we need a special slide, and special technician, so it [RDT] will be easier for that. RDT is a positive case, or the negative cases, and it's a simple method, so it will be useful and helpful to detect anything also.” | B1 |
|  | Lab technician | "So RDT, it is a simple test which can be performed everywhere. It takes less time and less skilled manpower because we needn't train much more on that. So, it makes things easier." | B2 |
| RDTs increase cholera screening | Physician, public lab | "From the past one year, we started having cholera rapid diagnostic test in our hospital and we came to realize that we have cholera here as well...Before that, severe patients who can't control loose stool were kept in the leaking bed and hanging drop test were done for such patients. Like now, when a patient comes with diarrhea, we think that it could be cholera. But before, we didn't think so." | C1 |
| RDTs for treatment | Physician, nonprofit hospital | "Early diagnosis, early treatment everything. No unnecessary test, no unnecessary management." | D1 |
|  | Physician, community hospital | "If he comes from a cholera-endemic area with severe AGE, with severe dehydration...before the investigation's report is coming to me, I have already started my treatment with cholera. That is the difference between clinician and epidemiologist or public health. We sometimes have to trust our clinical knowledge, intuitions, and the scenario which we first see the patient. We do send the test, and we do stop the treatment if it is wrong. But we might start treatment sometimes before our investigations comes." | D2 |
|  | Physician, public hospital | "RDT doesn't confirm cholera positive but still sometimes overtreatment is considered better than under treatment in medical science. If they have cholera, we need to give antibiotics. Rather than a patient dying from under treatment, sometimes for diarrhea cases, we over treat the patient. If we have started giving antibiotics to an RDT positive patient, we still continue with the antibiotics even if the culture report is negative." | D3 |
| RDTs in low-capacity settings | Head of Lab, private hospital | "That's very helpful because, locally, here we are in a tertiary center. Local person is more applicable because the primary hospital or the tertiary hospital is far from their homes. So, if you are using it, the number of detections, prevention and treatment of the patient will be easier, I think so." | E1 |
| RDT validity | Lab technician, private hospital | "And in case of a pandemic or if there is an outbreak, sometimes a false result will be high, because the manufacturing companies are all dealing with the profit and everything, and they don't look after the quality." | F1 |
| Limited awareness among physicians | Physician | "Because for me, if I have to order, I have to get literature about the comparable, how effective it is. You might be on a surveillance phase, but for us, as a treating physician, I should have complete information regarding what it is because you see, there are some cases where rapid antibiotics have been used unnecessarily in the patients." | G1 |
|  | Physician | "Since we have choices in Kathmandu valley and RDT is there, ELISA is there, CLIA is there even PCR is there. So, if there isn't an emergency condition, the doctors prefer ELISA or CLIA methods rather than RDT... RDT is used in emergency conditions when there is less time... But if the client has time, then, why don't we go to the higher technology? That's their logic. And I agree with that because the name itself is rapid diagnostic test. So, when we need our result rapidly, then, it's preferred.” | G2 |
| Lack of training | Physician, private hospital | “Because I think [untrained health workers] will not pick up the cases properly. They won’t go as per the case definition. In rural areas, everyone knows everybody, so, everybody would like to know whether they have cholera or not. If the public know that there is a test that can detect cholera, then, it influences the health worker to do it.” | H1 |
| Low prioritization and insufficient cholera infrastructure | Government official | “Up to now, I don't think the program has envisioned the cholera RDT as part of routine diagnosis. [For other diseases], yes, there is budget [for RDTs]. So, it is via government or maybe by EDP, but EDP will roll out through the government programs only for diseases like HIV, malaria, lymphatic filariasis, kalazar. So, plenty of the government budgets are there for the diseases of national importance, which are targeted for elimination or eradication. So, this is where we are relying on the RDTs... For cholera RDTs, if the surveillance shows a rise, it will be from the 118 sites. So, that will be enough evidence.” | I1 |
|  | Laboratory technician | “If there are many cases coming, then only media will be available; but such cases were not coming. As the media was not ready, we didn't do it here.” | I2 |
|  | Lab technician, private hospital | “The separate hospital system is there because our system is the one we are using. The hospital system software is there, so we record everything in the software.” | I3 |

## RESULTS

### Quantitative Results

#### EWARS monthly surveillance reports

In the publicly available national EWARS database, 8,859 acute gastrointestinal enteritis (AGE) and 11 cholera events were reported from 113 out of 118 sentinel sites from June through November 2023. Of the 8,870 AGE cases reported, 43.9% (3,892) indicated that no confirmatory laboratory test was conducted; 229 (2.6%) indicated that a cholera RDT was used.

During this period, RDTs were delivered to EWARS sites beginning mid-July and documented as received by the end of August. RDTs were delivered with printed job aids but in the 106 out of 114 EWARS sites not included in the IVI intensive surveillance research study, no additional training was included.

#### Monthly cholera RDT utilization survey at EWARS sites

Sixty-seven EWARS sites completed the RISE study REDCap survey at least one time from August to December 2023. Among the 67 reporting facilities, on average, facilities completed 2 monthly surveys (range: 1-6) One hospital completed the survey twice in one month. Of the 24 hospitals that completed the survey in December and thus reported on the full monitoring period from August-December 2023, a median of 0 RDTs were performed (range: 0-20). Of the 67 EWARS sites, 67.7% (44/67) reported never performing a cholera RDT and 20.0% (13/67) performed one to five RDTs prior to filling out the survey. A total of 141 RDTs were performed across all 67 reporting health facilities; a total of 5 positive RDTs were reported from four health facilities.

In the 44 facilities that reported zero RDT use, the most common reasons given for why RDTs were not used were that no suspected cholera cases arrived in their health facility (18/44; 41%) and that doctors did not order cholera RDTs (6/44; 14%). Many facilities did not provide a reason for not using RDTs (17/44; 39%).

#### Nested surveillance cholera RDT testing

A total of 656 stool samples from patients seen at the 12 RISE nested surveillance facilities were tested via cholera RDT. From June to December 2023, 97.6% (640) of RDTs were negative, 2.1% (14) of RDTs were positive, and 0.3% (2) RDTs were inconclusive.

### Qualitative Results

Thirty-three interviews were completed across the three target groups: 12 physicians with experience treating diarrhea patients within one of the RISE nested EWARS health facilities, 15 laboratory technicians with experience performing cholera microbiology tests within one of the RISE nested surveillance EWARS health facilities, and 6 government officials from the EDCD and NPHL. Key findings from the interviews are summarized by theme and exemplary quotes outlined in Table 2.

### Perceived Benefits of RDTs

#### RDTs facilitate cholera surveillance

Cholera RDTs were recognized by participants as beneficial for facilitating cholera surveillance and swift outbreak detection. All groups of participants said cholera RDTs enabled rapid, early detection of cases on site, given their ease of use and low cost. Thus, the RDTs were viewed as allowing for increased screening without putting undue burden on the health facility, making them scalable especially in outbreak situations [Table 2, A1].

Several governmental stakeholders also explained that when samples from suspected cholera cases need to be sent to reference laboratories, typically at the provincial and central level where there is the capacity to perform culture confirmation, there are often issues with tracking these samples. A government interviewee elaborated, explaining that when a health facility with the suspect case needs to send a sample, they first must request Cary Blair (CB) the preservation media, which takes one to three days on average to reach the site. Then, it usually takes 1-3 days for the preserved sample to return to the NPHL, and finally 1-2 days for confirmation tests to be performed. Cumulatively, from being identified as a suspect case based on clinical signs to confirmation is at best 3-8 days. The challenges associated with sending a sample to reference labs cause delays in testing, participants said, and when combined with the possibility that samples were preserved incorrectly there is worry about false results. These participants said that RDTs offer a way to better triage which samples need to be sent to reference laboratories and can resolve some of the timing issues by providing an early alert for outbreak response teams.

#### RDTs are easy to perform

Laboratory staff, the personnel primarily responsible for performing the RDTs, said that they are “easy enough” to be done by anyone. Participants believed RDTs would help reduce the burden on laboratories and ensure that the specialized technicians have more time to focus on other testing needs [Table 2, B1].

The RDTs were viewed by some technicians as being more accurate than the commonly performed hanging drop test. Many technicians also felt that the RDTs would be most useful for unskilled personnel in facilities with limited laboratory capacity, where other cholera diagnostic tests are unavailable and disease rates are typically higher in their experience [Table 2, B2].

#### RDTs increase cholera screening

Several participants noted that prior to receiving the RDTs, physicians did not often order cholera-specific lab tests, like culture, as they are time and cost intensive, and often delivered unreliable results. It was also believed by many physicians and lab technicians that cholera was not endemic in Nepal. Having the RDTs available increased screening, surprising participants when they realized cholera was still circulating in their communities. It was reported by lab interviewees that the tests not only increased awareness of cholera but were also cost-effective and less time-intensive compared to traditional laboratory methods [Table 2, C1].

In addition, government officials recognized broader advantages of national RDT deployment, emphasizing their efficiency in detecting cases, generating trackable data, and their suitability for resource-limited settings.

#### RDTs for treatment

Participants, primarily physicians, viewed cholera RDTs as an opportunity for faster diagnosis and treatment. Few physician participants understood that the RDT is not intended to be used for case management [Table 2, D1].

Several interviewees stated that an RDT positive combined with clinical suspicion would be enough to start treatment with antibiotics. They felt that due to the rapid deterioration of patients with cholera, it is better to over-treat than under-treat patients. Some clinicians mentioned stopping the antibiotics when a culture comes back negative, while others say they continue with the full antibiotic course regardless [Table 2, D2].

While some physicians say they would rather not prescribe antibiotics without a lab report confirming they are needed, delays in testing make it so they often will have to skip testing and administer antibiotics based on their clinical assessment. Some physicians said cholera RDTs could allow them to make better informed decisions about antibiotics and thus limit misuse of antibiotics. However, it is noted by participants that if RDT results are not received prior to the initiation of treatment then they will have no impact [Table 2, D3].

#### RDTs in low-capacity settings

Many participants mentioned that using RDTs can prevent unnecessary patient travel. For example, for patients living in rural parts of the country, who would need to travel to or have samples sent to high-capacity laboratories, the cholera RDTs can save time and money by preventing unnecessary confirmatory testing. Many participants felt strongly that the RDTs should be distributed beyond the EWARS network to include health facilities at smaller, more local levels; by having RDTs in health posts, it would allow doctors to “intervene with the patient instantly” and treatment could occur locally [Table 2, E1].

### Perceived negatives of RDTs

#### RDT validity

Negative perceptions of the sensitivity and specificity of the cholera RDTs was a recurring theme across the lab technicians, physicians, and government officials interviewed. Although government officials acknowledged the potential advantages of cholera RDTs, they expressed ongoing concerns regarding their reliability and accuracy. They said that while cholera RDTs have the potential to become the “test of choice”, it must be confirmed that they are of high manufacturing quality and validity [Table 2, F1].

For physicians, the perceived low validity of the cholera RDTs negatively impacted their acceptance of RDTs. Many physicians said that they needed to understand the validity of the test before considering it as part of their cholera workup. These concerns were illustrated by participants through examples of events where a positive RDT result was disproven by a negative culture report. In these cases, the RDT result caused unnecessary panic for the patients and made the physicians realize that they were treating “in the line of cholera” for patients that did not need it. Amongst physicians that had never used the cholera RDTs before, their experience with COVID RDTs and with rapid HIV tests led them to doubt that cholera RDTs would provide reliable results. Laboratory technicians also expressed concerns about false positives, but many of them stated that false positives could occur not only due to poor manufacturing and quality of the kit, but also from kits being exposed to extreme temperatures or getting wet, inadequate stool collection, or failure to wait the appropriate amount of time to read the results.

Several government and physician interviewees said that since there are no WHO pre-qualified cholera RDTs available, they do not feel comfortable recommending the test. Lab technicians agreed, explaining that the lack of pre-qualified cholera RDTs available was concerning and that WHO recommended assays and equipment are superior and received most favorably by physicians since Nepal is not able to perform their own quality checks.

#### Cost and accessibility

Several physicians said that they would choose the cheapest diagnostic option available, and if necessary, would rather treat without laboratory testing if it saved their patient money. Physicians were wary about ordering RDTs for their patients given uncertainty about whether RDTs would be consistently available. Although most participants recognized potential benefits from RDT use, they highlighted that these benefits could only be realized if RDTs were consistently available, physicians were adequately trained on prescribing the RDTs, and if the results are reported systematically.

### Challenges with implementing RDTs

#### Limited awareness among physicians

Stakeholders noted that physicians were a key barrier to uptake of cholera RDTs at the facility level given that physicians must order or prescribe the cholera RDT before a lab technician can perform it. About half of the physicians interviewed were not yet aware that cholera RDTs were available at their facility. Among physicians who were aware of the cholera RDTs, some considered it to be the preferred lab test for cholera, while others hesitated to order RDTs given doubts about validity, as mentioned previously [Table 2, G1].

A few laboratory technicians noted that physicians continued to order hanging drop (motility) tests given lack of awareness about the cholera RDTs. Physicians described the motility test as a “rapid test” given that it can be completed in a quarter of the time as a culture, which was described as taking around 12 hours to complete. Some laboratory technicians mentioned there were no mechanisms for ensuring that all staff knew about the cholera RDTs following the arrival of the first shipment, and in some cases, the technicians themselves spoke to the doctors directly to encourage them to start ordering the tests.

A minority of physicians expressed a preference for the cholera RDTs given hanging drop is not recommended by the WHO; they explained that the subjectivity, expertise, and manpower required to perform hanging drop makes them believe that RDTs are better. However, nearly half of the physicians interviewed cited hanging drop as their preferred test. Many believed the sensitivity and specificity are higher for hanging drop and reasoned that they have been using hanging drop for a long time without issues. These physicians said they would need to see literature proving that the RDTs have higher validity than hanging drop to consider changing their perspective [Table 2, G2].

#### Lack of training

Almost all interviewees (both laboratory technicians and physicians) expressed a desire for more training on cholera RDTs. According to interviewed laboratory technicians, limited uptake of the RDTs was likely due to a lack of training for physicians. They explained that without training, physicians likely did not know when and for whom to order a cholera RDT, the definition of a cholera outbreak, or how to handle a stool sample for testing. Lab technicians also expressed that in the lab, inadequate training can lead to false positives when the tests are not done correctly. They explained, for example, failing to wait the appropriate amount of time before reading the test or failing to adequately instruct the patient on proper stool collection techniques can result in false results. Another mentioned how a lack of training can lead to misuse or providing the RDT to people who do not necessarily meet the case definition [Table 2, H1].

In addition, interviewees described varying methods for recording cholera cases given that each site had differing internal systems. Several laboratory technicians stated that laboratory results are sent to the EDCD and EWARS through their facility’s medical reporter, not by the laboratory. Both lab technicians and physicians felt consistent training would be necessary to ensure uptake of the RDTs and accurate reporting to maximize their impact. Interviewees offered recommendations for cholera RDT training (Table 3).

**Table 3.**
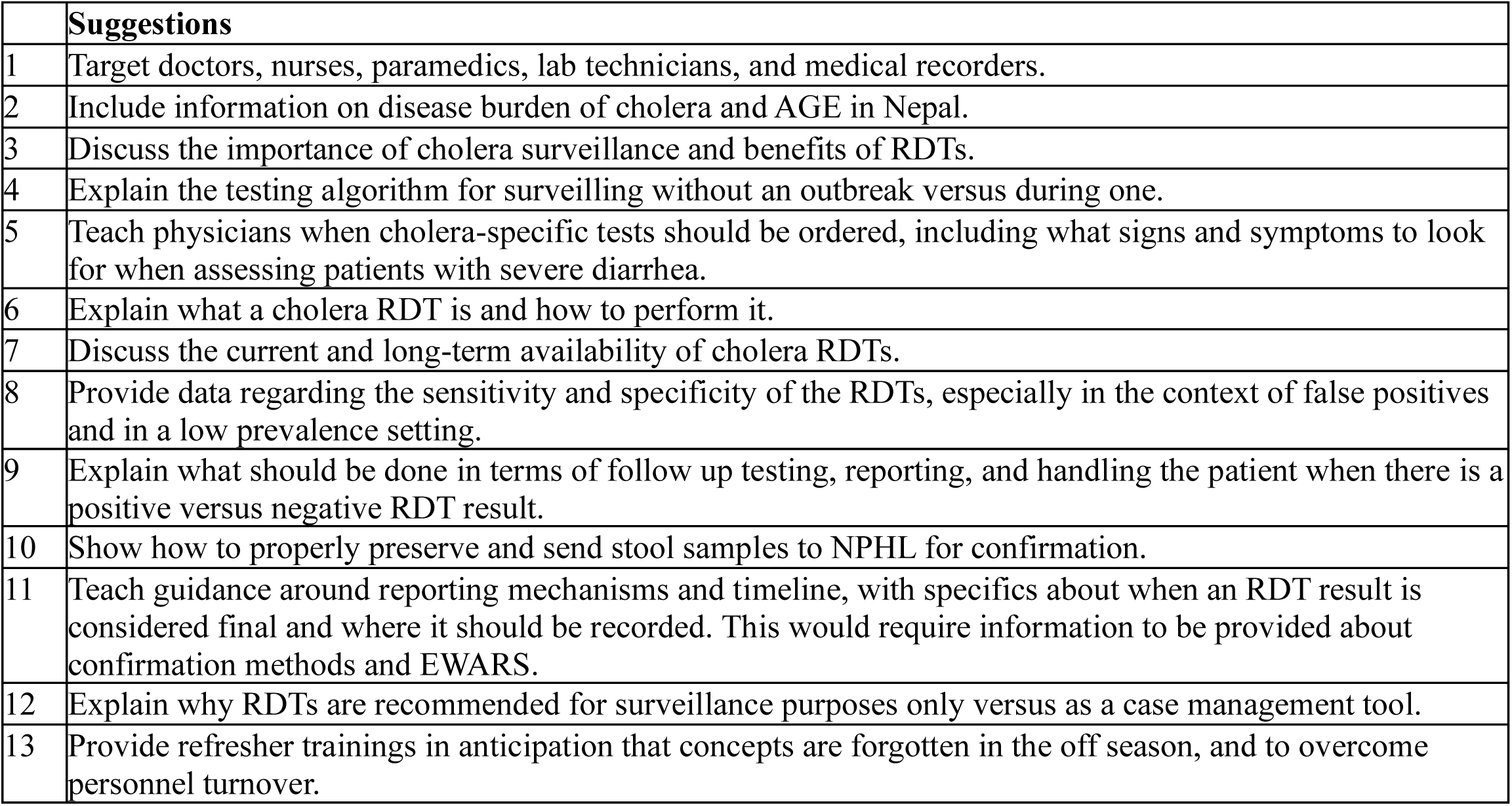
Interviewee suggestions for training on cholera RDTs.

|  | **Suggestions** |
| --- | --- |
| 1 | Target doctors, nurses, paramedics, lab technicians, and medical recorders. |
| 2 | Include information on disease burden of cholera and AGE in Nepal. |
| 3 | Discuss the importance of cholera surveillance and benefits of RDTs. |
| 4 | Explain the testing algorithm for surveilling without an outbreak versus during one. |
| 5 | Teach physicians when cholera-specific tests should be ordered, including what signs and symptoms to look for when assessing patients with severe diarrhea. |
| 6 | Explain what a cholera RDT is and how to perform it. |
| 7 | Discuss the current and long-term availability of cholera RDTs. |
| 8 | Provide data regarding the sensitivity and specificity of the RDTs, especially in the context of false positives and in a low prevalence setting. |
| 9 | Explain what should be done in terms of follow up testing, reporting, and handling the patient when there is a positive versus negative RDT result. |
| 10 | Show how to properly preserve and send stool samples to NPHL for confirmation. |
| 11 | Teach guidance around reporting mechanisms and timeline, with specifics about when an RDT result is considered final and where it should be recorded. This would require information to be provided about confirmation methods and EWARS. |
| 12 | Explain why RDTs are recommended for surveillance purposes only versus as a case management tool. |
| 13 | Provide refresher trainings in anticipation that concepts are forgotten in the off season, and to overcome personnel turnover. |

#### Low prioritization and insufficient cholera infrastructure

Some government officials were skeptical about the importance of cholera RDTs given the perceived low prevalence of cholera in the country. One government stakeholder stated that in Nepal, there is limited investment in cholera as it is not seen as a “disease of national importance.” They provided the example of malaria, where because there is a national commitment to eliminate malaria by 2026, malaria RDTs are being widely distributed and utilized to reach the testing minimum set by the government. Another interviewee agreed, stating that they do not believe cholera RDT distribution will be sustainable beyond this pilot study because cholera is low on the list of priorities for the government, so money and manpower will likely not be spared for surveillance enhancement [Table 2, I1].

Across all groups, many interviewees stated the need for increased cholera diagnostic capacity to be able to perform culture confirmation for samples that tested positive by RDT, including options that have a long shelf life such as filter paper, given difficulties with transport during cholera season [18]. A laboratory technician highlighted that in many cases, while facilities may technically have culture capability, this does not necessarily mean having cholera culture capacity. In many sites, necessary lab supplies are not usually on hand unless there is a clear need [Table 2, I2].

Many samples need to be transported elsewhere for confirmation testing, to labs like the NPHL, causing delays in case confirmation and reporting. Government officials confirmed that EWARS has significant gaps in data, as sites frequently do not submit their reports on time or at all, since many facilities either preferred their internal system or were not aware of EWARS [Table 2, I3].

## DISCUSSION

This evaluation uncovered several critical factors regarding deployment and use of RDTs to improve cholera control and elimination. Though participants expressed positive perceptions of cholera RDTs, doubts about the RDTs’ validity and availability were pervasive. Additionally, physicians wanted to use RDTs to guide case management, though RDTs were not intended for that purpose. These factors, coupled with limited awareness and training in the correct usage and interpretation of cholera RDTs, likely constrained RDT adoption, as documented in the case of malaria RDTs [19].

Interviewees from all groups in the RISE study highlighted the potential utility of RDTs beyond Kathmandu and EWARS. Other studies in Kathmandu Valley [7] and elsewhere [19, 20, 21, 22] have similarly highlighted the need for expanding RDTs to improve disease surveillance and response [4]. However, some participants felt that distributing cholera RDTs beyond the designated EWARS facilities in Nepal could lead to misuse, particularly overuse, of the RDTs given the lack of public health oversight in these areas. These concerns are supported by published data reporting frequent misuse of malaria RDTs during implementation [23]. In these cases, authors stressed the importance of providing clear guidelines for use through training with various healthcare workers and community members [23].

The most common concern participants expressed was the belief that RDTs yielded a high degree of false-positive results. Stakeholders primarily focused on how false positive RDTs resulted in unnecessary isolation and antibiotic treatment. Culture, which is the typical confirmation test for suspected cholera cases in Nepal, is recommended only for samples which are RDT positive. However, misconceptions about validity of RDT results led a number of lab technicians to perform confirmatory tests for all samples, meaning that cholera RDTs were adding to their workload rather than increasing efficiency. In Nepal, hanging drop is often the method of choice for screening suspect cholera samples, though it is not recommended by the Global Task Force on Cholera Control (GTFCC) due to the expertise required [24]. These findings suggest the need for public health officials to provide information to physicians about RDT performance, and the strengths of this test compared to hanging drop. Physician awareness and acceptance of the rapid tests is crucial for their utilization [22].

The recent GTFCC Cholera Surveillance Guidance recommends RDT use at surveillance units to rule out cholera, achieve early detection, and monitor outbreak trends [5, 11, 24]. Given that existing literature finds an over-reporting of cholera during outbreaks when cases are identified solely based on clinical signs and symptoms without laboratory confirmation [4], RDT use can reduce over-reporting and unnecessary confirmatory testing, saving money and time [1, 8, 9, 10]. It is important to note that the quantitative data, namely the nested and monthly REDCap surveys, from 2023 of the RISE project was not representative of the total amount of RDTs performed in Nepal and did not include national confirmation data. This paper is a reflection of only the data received through the RISE surveys and not inclusive of the intensive surveillance data, which will be submitted for publication in quarter two of 2025.

To encourage health workers to integrate the cholera RDT into their routine practice, interviewees emphasized the need for assured program longevity. A clear explanation of the Gavi platform funding through 2025, and potential expansion through 2030, should be conveyed to relevant personnel to ensure that cholera RDT utilization is not hampered by concerns of future availability, quality, and cost. Local government stakeholders should be involved in the development of project plans and monitoring practices to ensure sustainability and country-level procurement, as has been found successful for malaria RDT scale-up efforts [23].

Another commonly reported barrier to RDT implementation was the lack of knowledge and training to use and report cholera RDTs correctly. Other studies cited similar challenges around RDT implementation (including misconceptions about their intended use, accuracy, and physician uptake barriers) and reporting [7, 19, 20, 22, 23]. Communication, training, and sensitization to the tests and their purpose were deemed crucial and often done prior to their distribution to emphasize their benefits and minimize negative perceptions [19, 20, 22, 23]. Comprehensive training and reinforcement for both physicians and laboratory technicians, both on RDT usage and reporting, should be incorporated into national platforms, with support from Gavi, GTFCC, and other global experts. Improving reporting is crucial for generating interpretable and informative data, and for strengthening surveillance and the probability of future support for RDTs and OCV through external donors.

Finally, when describing the benefits of RDTs, physicians focused on ways they could utilize the cholera RDTs to aid in their case management. However, this contradicts the WHO recommendation that RDTs be used for surveillance purposes only, not diagnostic. The overlap with case management versus surveillance may, in part, be due to clinicians having extensive experience with malaria RDTs, which are used in a case management capacity to prescribe artemisinin-combination therapy (ACT) [19, 23]. This highlights a need for public health officials to articulate the usefulness of the results and how they should be applied in line with WHO and GTFCC recommendations.

### Strengths and Limitations

Our study is the first to obtain an in-depth understanding of cholera RDT use through a robust quantitative and qualitative analysis. We sought to achieve credibility through triangulation (multiple types of participants and qualitative and quantitative data) and peer debriefing. We aim to enhance transferability by providing thick descriptions of the research context and participants, and purposeful sampling to ensure we obtained the most relevant and informative information. Confirmability was promoted through maintaining an audit trail and continuous reflexivity to expose and address researcher bias. We also pursued dependability through an iterative coding process in which multiple researchers revisited and reanalyzed data to check for consistency [27].

Importantly, at the time of data collection for Year 1 (2023) of the RISE project, the application portal for Gavi’s Diagnostic Procurement Platform was opened for eligible countries. For countries that submitted applications in 2023, cholera RDTs were scheduled to arrive between July 2024 and February 2025. The Nepal government applied for a supply of over 40,000 RDTs in anticipation of the 2024 monsoon season and their application was approved. Findings from Year 1 of the RISE project were shared with Nepali government stakeholders improving the country’s preparedness for large-scale roll-out of cholera RDTs.

Some limitations should also be noted. Though we aimed to ensure all levels of personnel involved in cholera surveillance were included in this study, most participants were higher-level as many of the lower-level technicians or physicians not in charge of their departments were not available or comfortable to be interviewed. This may result in some gaps in our understanding of both the in-country perspective and some additional barriers to successful cholera surveillance execution. The findings and experiences reflected in this paper illustrates the first year of using cholera RDTs on a wide scale, in Nepal and globally, and it may, in time, help establish cholera RDTs as a routine method and integration of the tool into standard practice. We expect that attitudes are likely to change over time. We will aim to capture and expand on such attitudinal changes in the upcoming RISE Year 2 reports.

All the interviews were conducted with participants in the capital city of Kathmandu; perspectives and experiences from personnel in more rural areas of the country may differ. Additionally, interviews were conducted shortly after cholera RDTs were delivered to some facilities; thus, some participants had limited or no direct experience with the RDTs prior to the interview. However, these participants still provided important information about how cholera RDTs were viewed at the point of their introduction into facilities. Despite these limitations, our study provides a unique understanding of the barriers and facilitators to cholera RDT implementation.

## CONCLUSION

Despite the many recognized benefits of cholera RDTs, challenges and misconceptions persist. Addressing these barriers to successfully scale-up cholera RDTs in Nepal and beyond requires ensuring a consistent and reliable supply chain, maintaining strict quality control, addressing cost-effectiveness and quality concerns, and gradual expansion to non-centralized sites with robust monitoring. Also, clear guidelines and enhanced training on cholera RDT availability, administration, interpretation, reporting, and response to positive results are needed. Understanding existing national surveillance systems and cholera lab diagnostic needs prior to integrating cholera RDTs can be helpful for ensuring that distribution plans adequately address any potential issues that may arise. Though cholera prevalence in Nepal is low, a National Cholera Control Plan that incorporates the lessons from this analysis is essential to achieve surveillance targets and move towards the elimination of cholera.

## Data Availability

The de-identified data that support the findings of this study are available from the corresponding author upon reasonable request.

## Acknowledgments

We thank the staff of the Group for Technical Assistance (GTA) Foundation, Kathmandu, Nepal for their participation in this study. We also thank the International Vaccine Institute (IVI), including Derick Kimathi, Yubin Lee, and Haeun Cho, for their support in this project. Additionally, we want to extend our thanks to Nepal’s Epidemiology and Disease Control Division (EDCD), National Public Health Laboratory (NPHL), and Early Warning and Reporting System (EWARS) sites for their continued support and participation in this study.

## Funding statement

This study was supported by Gavi, The Vaccine Alliance and administered through the Johns Hopkins Bloomberg School of Public Health.

## Competing interests

No competing interests to declare.

## Disclaimer

The funder had no role in study design, data collection and analysis, decision to publish, or preparation of the manuscript.

## Notes

### Competing Interest Statement

The authors have declared no competing interest.

### Author Declarations

The Institutional Review Board (IRB) of The Johns Hopkins University Bloomberg School of Public Health gave ethical approval for this work.

