## Supplemental Tables for "Enhancing national cholera surveillance using rapid diagnostic tests (RDTs): a mixed methods evaluation"

### Supplemental Materials

**Table 1: Open-coding Framework sub-theme definitions for Government interviews.**

| | |
|---|---|
| A. Job description | Unlike with the lab and physician transcripts, government stakeholders will have varied roles. While this information is not a "theme" and will not be analyzed thematically, it will be used for understanding of the background/context from which the interview subjects are speaking. Record brief but relevant description of subject's job/role especially as it relates to cholera surveillance. |
| B. Barriers | Record any details on subjects' perspective on critical barriers to the implementation of RDT surveillance at sites. Examples may include perspectives on addition of cholera RDTs to cholera testing and surveillance protocols/algorithms, critical barriers to RDT implementation at sites, degree to which facility-level activities like ordering tests or reporting results are siloed vs. integrated, disincentives to perform/interpret/order tests, availability of tests or physicians or lab personnel etc. |
| C. Facilitators | Record any details on subjects' perspective on critical success factors to the implementation of RDT surveillance at sites. Examples may include perspectives on addition of cholera RDTs to cholera testing and surveillance protocols/algorithms, critical facilitators to RDT implementation at sites, degree to which facility-level activities like ordering tests or reporting results are siloed vs. integrated, incentives to perform/interpret/order tests, availability of tests or physicians or lab personnel. |
| D. Fidelity and Fit | Record any details on subjects' perspective on: 1) If integration and ordering of cholera RDTs at surveillance sites were implemented as intended; 2) How and why changes were made to the implementation strategy in response to the context. Examples could include how/if any material (i.e., resources, time, workload, etc.), cultural (i.e., work environment, interpersonal/ partnership/stakeholder/implementer dynamics etc.) or contextual factors (i.e., epidemiological changes, politics etc.) external to RISE impacted RDT implementation; 3) If/How RDT implementation strategies , distribution, training and data integration (i.e., printed aids, PPHL meeting) were appropriate/effective for Nepali cholera surveillance and detection. |
| E. Nepali Cholera Surveillance System - Structure and Reporting | Record any information shared by subject on the structure of the Nepali infectious disease surveillance system, or how information is reported within that system. We are primarily interested in the system as it relates to cholera, but also include relevant comparisons/references made to other disease surveillance/reporting mechanisms. Examples could include surveillance or reporting guidelines or processes (e.g., steps of outbreak declaration), major infrastructure, procedures or hierarchies of different entities involved in surveillance activities, modes of reporting (e.g., electronic, by phone), differences by geographic region or facility type etc. |
| F. Lab Diagnostics for Pathogen Detection/Confirmation OTHER than the Cholera RDT | Record any information shared by subject on procedures or methods of testing pathogens for confirmation in the context of surveillance - other than cholera RDTs. We are primarily interested in detection/confirmation as it relates to cholera, but also include relevant comparisons/references made to other pathogens. Examples may include types of tests performed (e.g., Cary Blair, PCR, serology) and why certain methods are preferred, challenges/advantages of different methods, logistics associated with confirmation processes etc. |
| G. Roles in cholera surveillance | Record any details on subjects' perspectives on their roles or the roles of others in cholera surveillance in Nepal. |
| H. POV on Negatives of RDT Use | Include any information on subjects' thoughts on the challenges/limitations/drawbacks of using cholera RDTs in Nepal. Examples could include implications for screening, surveillance and disease control, workload, reporting, logistics, relative barriers for |

| | |
|---|---|
| | different facility types or geographical areas, seasonality, private vs. public sector factors, cost to patients/government, specificity and sensitivity, patient care etc. |
| I. POV on Benefits of RDT Use | Include any information on subjects' thoughts on the benefits of using cholera RDTs in Nepal. Examples could include implications for screening, surveillance and disease control, workload, reporting, logistics, relative facilitators for different facility types or geographical areas, seasonality, private vs. public sector factors, cost to patients/government, specificity and sensitivity, patient care etc. |
| J. Additional Recommendations | Record information on additional recommendations subjects have about cholera RDT use in Nepal. Examples could include suggestions for monitoring and evaluation, edits to cholera outbreak protocols/algorithms, training, rollout and scale-up, government oversight, distribution, job aids, sensitization etc. |
| K. Other content, Comments, Questions, Follow-ups | Record information that doesn't fit into the other columns, but seems important/relevant to the overall study, questions you may have that need to be followed-up on, suggestions from subjects that should be followed-up on etc. |

**Table 2: Open-coding Framework sub-theme definitions for Physician interviews.**

| | |
|---|---|
| A. Barriers | Record any details on 1) Critical barriers to RDT implementation at surveillance sites. Examples could include degree to which facility-level activities like ordering tests or reporting results are siloed vs. integrated, disincentives to perform/interpret tests or disinterest, availability of tests or lab personnel. |
| B. Facilitators | Record any details on 1) Critical success factors to RDT implementation at surveillance sites. Examples could include degree to which facility-level activities like ordering tests or receiving results are integrated vs. siloed, availability of tests or lab personnel. |
| C. Fidelity and Fit | Record any details on 1) If integration and ordering of cholera RDTs at surveillance sites were implemented as intended; 2) How and why changes were made to the implementation strategy in response to the context. Examples could include how/if any material (i.e., resources, time, workload, etc.), cultural (i.e., work environment, interpersonal/ partnership/stakeholder/implementer dynamics etc.) or contextual factors (i.e., epidemiological changes, politics etc.) external to RISE impacted RDT implementation; 3) If/How RDT implementation strategies , distribution, training and data integration (i.e., printed aids, PPHL meeting) were appropriate/effective for Nepali cholera surveillance and detection. |
| D. Experiences performing cholera RDTs | Record any details on physicians’ experiences with the process of ordering cholera RDTs or the process of receiving test results. Examples could include their thoughts on ordering protocols, process of ordering a cholera RDT including as it relates to patient interactions (i.e., any forms to complete, patient counseling, getting a sample etc.). |
| E. Experiences diagnosing cholera | Record any details on physicians’ experiences diagnosing cholera. Examples could include their thoughts on symptoms, decision-making process as it relates to of diagnosis vs. treatment and/or surveillance, utility of or reasons/motivations to diagnose, options for diagnosis as it relates to different types of tests and reasons for selecting one over another, decision-making process as it relates to deciding if it’s a suspect cholera case and the decision to test or treat based on that determination. |
| F. Roles in cholera surveillance | Record any details on physicians’ perspectives on their roles or the roles of others in cholera surveillance in Nepal. |
| G. POV on Negatives of RDT Use | Include any information physicians' thoughts on the challenges/limitations/drawbacks of using cholera RDTs in Nepal. Examples could include implications for screening, surveillance and disease control, workload, reporting, logistics, relative barriers for different facility types or geographical areas, seasonality, private vs. public sector factors, cost to patients/government, specificity and sensitivity, patient care etc. |
| H. POV on Benefits of RDT Use | Include any information physicians' thoughts on the benefits of using cholera RDTs in Nepal. Examples could include implications for screening, surveillance and disease control, |

| | |
|---|---|
| | workload, reporting, logistics, relative benefits to different facility types or geographical areas, seasonality, cost to patients/government, specificity and sensitivity, patient care etc. |
| I. Additional Recommendations | Record information on additional recommendations physicians have about cholera RDT use in Nepal. Examples could include suggestions for training, rollout and scale-up, government oversight, distribution, job aids, sensitization etc. |
| J. Other content, Comments, Questions, Follow-ups | Record information that doesn't fit into the other columns, but seems important/relevant to the overall study, questions you may have that need to be followed-up on, suggestions from subjects that should be followed-up on etc. |

**Table 3: Open-coding Framework sub-theme definitions for Laboratory Technician interviews.**

| | |
|---|---|
| A. Barriers | Record any details on 1) Critical barriers to RDT implementation at surveillance sites. Examples could include degree to which facility-level activities like reporting results are siloed vs. integrated, disincentives to perform/interpret tests or disinterest, availability of tests. |
| B. Facilitators | Record any details on 1) Critical success factors to RDT implementation at surveillance sites. Examples could include degree to which facility-level activities like reporting results are integrated vs. siloed, availability of tests. |
| C. Fidelity and Fit | Record any details on 1) If distribution and integration of cholera RDTs at surveillance sites were implemented as intended; 2) How and why changes were made to the implementation strategy in response to the context. Examples could include how/if any material (i.e., resources, time, workload, etc.), cultural (i.e., work environment, interpersonal/ partnership/stakeholder/implementer dynamics etc.) or contextual factors (i.e., epidemiological changes, politics etc.) external to RISE impacted RDT implementation; 3) If/How RDT implementation strategies , distribution, training and data integration (i.e., printed aids, PPHL meeting) were appropriate/effective for Nepali cholera surveillance and detection. |
| D. Experiences performing cholera RDTs | Record any details on laboratorians' experiences performing cholera RDTs. Examples could include their thoughts on testing protocols, ease of use (may be relative to other methods of testing), what would make the tests easier and more convenient to perform etc. |
| E. Experiences interpreting cholera RDTs | Record any details on laboratorians' experiences interpreting cholera RDTs. Examples could include their thoughts on reading test results (may be relative to other methods of testing), false positives, diagnosis, application to outbreak algorithms, what would make the tests easier and more convenient to interpret etc. |
| F. Roles in cholera surveillance | Record any details on laboratorians' perspectives on their roles or the roles of others in cholera surveillance in Nepal. |
| G. POV on Negatives of RDT Use | Include any information laboratorians thoughts on the challenges/limitations/drawbacks of using cholera RDTs in Nepal. Examples could include implications for screening, surveillance and disease control, workload, reporting, logistics, relative barriers for different facility types or geographical areas, seasonality, private vs. public sector factors, cost to patients/government, specificity and sensitivity etc. |
| H. POV on Benefits of RDT Use | Include any information laboratorians thoughts on the benefits of using cholera RDTs in Nepal. Examples could include implications for screening, surveillance and disease control, workload, reporting, logistics, relative benefits to different facility types or geographical areas, seasonality, cost to patients/government, specificity and sensitivity etc. |
| I. Additional Recommendations | Record information on additional recommendations laboratorians have about cholera RDT use in Nepal. Examples could include suggestions for training, rollout and scale-up, government oversight, distribution, job aids, sensitization etc. |
| J. Other content, Comments, Questions, Follow-ups | Record information that doesn't fit into the other columns, but seems important/relevant to the overall study, questions you may have that need to be followed-up on, suggestions from subjects that should be followed-up on etc. |
